# Frequency of ocular manifestations in rheumatic autoimmune diseases (ARDs); a cross-sectional study

**DOI:** 10.1101/2024.01.08.24300887

**Authors:** Usama Sohail

**Affiliations:** Shaikh Zayed Hospital

## Abstract

Autoimmune disorders involve diverse symptomatic manifestations constituting an abnormal or exaggerated immune function and serve to attack self-antigens and result in tissue destruction. This study aims to comprehensively analyse the prevalence, type, associative factors, and the resultant impact of ocular symptoms on patients presented to rheumatology departments of multi-center tertiary care hospitals in Lahore. Methods. A total of 300 patients were studied during the time frame of Jan 2021-Dec 2022. Results. Rheumatoid Arthritis comprised 45.7 % (n=137) of patients, Systemic Lupus Erythematosus (SLE) was present in 34.3% (N=103) patients. Spondylarthritis 8.3%, Granulomatous Polyangiitis 3.3%, Juvenile idiopathic arthritis 0.7%, and Bechet Disease 2.3% were present. The total number of patients who developed eye manifestations was 18%. Keratoconjunctivitis Sicca was observed in 4 females and 2 males with overall frequency of 11.1%. Conclusions. Our study showed a surge in the cases of SLE diagnosed and witnessed during the period of 2022, but our study showed an overall decrease in the incidence of eye complications.

## INTRODUCTION

Autoimmune disorders involve diverse symptomatic manifestations constituting an abnormal or exaggerated immune function and serve to attack self-antigens and result in tissue destruction. (1) 80 different types of autoimmune disorders have been recognized. (2) A statistic from the United States of America showed an increased incidence of autoimmune disorders, around 5-7%, with females being the major targeted population. (3) Diseases with positive “Rheumatic Factor” are the main focus for the associated extra-articular effects and include a broad-spectrum range, including Bechet Disease, Systemic Lupus Erythematosus (SLE), Rheumatic Arthritis (RA), Dermatomyositis (DM), Vasculitis, Sjogren’s Syndrome and Polymyositis (PM). (4) Autoimmune Rheumatic Diseases (ARDs) target people globally, with a count affecting millions of people. Worldwide prevalence of SLE, RA, DM/PM, SS, Sjogren’s Syndrome and are 0.02–0.07% (5), (6), 0.5–1.1% (7), 0.02% (8), 0.02– 0.04%, (9) 0.05–4.8%, (10) respectively. ARDs can involve many body organs outside the joints, including skin, heart, lungs, eyes, kidneys, etc. Our research purpose is to find a link between rheumatic autoimmune disease and their ocular manifestations (11). Ocular symptoms containing autoimmune diseases mainly consist of Rheumatoid Arthritis (12), Sjogren Syndrome (13), Spondylarthritis (14), (15), Juvenile Idiopathic Arthritis, Systemic Sclerosis Granulomatosis with Polyangiitis (16), (17), Systemic Lupus Erythematosus associated with antiphospholipid syndrome (APA), Relapsing Polychondritis. (18) Inclusion of different eye tissues in patients with rheumatic diseases can lead to troublesome effects, of which the most distinctive ones are retinal vasculitis, silica, keratoconjunctivitis, glaucoma, and uveitis. The delicacy of the eye (19) as an organ can be indicated by the clinical signs that appear as the first indicator of any systemic distortion that leads to a change in its sensitive microenvironment. (20) A specialized sterile environment is present between blood and eye compartments (containing aqueous and vitreous humor), and rheumatic autoimmune disease tends to disturb this equilibrium. (21) But ocular problems following rheumatic diseases can present during the active stage or even years after the initial diagnosis. The optimal and gold standard treatment regimens for RA diseases have been disease-modifying anti-rheumatic drugs or DMARDs, as they are involved in halting the long-term progression of the disease and the resultant joint manifestations. (22) But the downside to these drugs is the unpremeditated ocular toxicity, especially those caused by the prototypic agent Methotrexate, retinal cotton-wool spots, orbital non-Hodgkin’s lymphoma, and ischemic neuropathy. (23) A study conducted in Mexico included 276 medical records in which the most common autoimmune rheumatic disease and ocular manifestation were spondylarthritis and keratoconjunctivitis sicca, respectively. (24) Another one done in a tertiary care setting in Lahore (Fatima Memorial Hospital) included 83 patients, in whom commonly occurring diseases were spondylarthritis and uveitis. (25) Hence, a detailed collaboration between the departments of rheumatology and ophthalmology is required for the ideal management of such patients. At best, routine-based checkups should be opted for as a planning methodology for diagnosing and treating such patients. As many diseases mentioned in this study result in acute visual complications, regular screening should be emphasized, even for asymptomatic patients. Till now, one such study has been conducted about this inter-relationship in our population, but more data is needed. This research aims to comprehensively analyze the prevalence, type, associative factors, and the resultant impact of ocular symptoms on patients presented to rheumatology departments of multi-center tertiary care hospitals in Lahore.

## MATERIAL AND METHODS

- Our study was retrospective cross sectional, carried out at Shaikh Zayed Hospital, Lahore. Data was taken between the time of Jan 2021 - Dec 2022. Data was included by probability method of simple random sampling. Out of total, a sample of 300 was taken. Sample included only those patients with autoimmune rheumatic diseases (RA factor +) and those who were diagnosed at the time by the rheumatologist. Our study was approved by the ethical review board and informed consent was taken to perform this study. Parameters assessed were demographics, presence of autoimmune disease and the resultant diagnosis of the eye disease. It also included the duration of the eye symptoms and the use of medication like steroids and the effect of medications.
- The data taken was analyzed by SPSS tool. Continuous variables like age were presented and the resultant frequency (percentage) was taken.

### Results

- A total of 300 patients were studied during the time frame of Jan 2021-Dec 2022. Average age ______ is taken. About 59.7% of the total data taken were females.
- Rheumatoid Arthritis comprised 45.7 % (n=137) of patients, SLE was present in 34.3% (n=103) patients. While Systemic Sclerosis 5.3% (n=16), Spondylarthritis 8.3% (n=25), Granulomatous Polyangiitis 3.3% (n=10), Juvenile idiopathic arthritis 0.7% (n=2) and Bechet Disease was present in 2.3% (n=7). The total number of patients who developed ocular manifestations was 18%. The individual ocular sub types of presents include, Keratoconjunctivitis Sicca 6% (n=18), Uveitis 5.3% (n=16), Scleritis 4% (n=12), and Peripheral ulcer keratitis 2% (n=6).

## DISCUSSION

Ophthalmological involvement is the main stay process we see in most autoimmune rheumatic processes. This study provides an overlook about the major eye complications in patients of Autoimmune Rheumatic Diseases. Most of the patients included in our study were cases of Uveitis and Keratoconjunctivitis Sicca.

### Autoimmune Rheumatic Diseases

During the time of our study (Jan 2021-Dec 2022). We took a total of 300 patients were taken. The data we had taken included the age between 36-45 years as the most prevalent age group. RA is the most common articular manifestation noticed in our study. It included 87 females and 50 males. Rheumatoid Arthritis was the most common prevalence seen during the period of 2021, but our retrospective study showed a surge in the cases of SLE diagnosed and witnessed during the time of 2022. Going further SLE was our second most common disease with a prevalence of 69 among females and 39 among males. Spondylarthritis was present in 10 females and 15 males. In reference to this study, one conducted in Fatima Memorial Hospital, Lahore demonstrated Spondylarthritis as the most common disease with the overall frequency of 38.6%. Out of 16 patients of SS, 10 females and 6 males were present. And lastly, GPA had a frequency of 4 males and 6 females.

### Ocular Manifestations

They constitute the most common extra-articular symptomology studied in autoimmune rheumatic spectrum. Our study included most of the cases including Uveitis and Keratoconjunctivitis Sicca as the ocular diseases. Keratoconjunctivitis sicca, also known as Dry syndrome, the most common etiological factor for which is Sjogren syndrome. In our clinical cases, diagnosis of KS is made by Schirmer’s test with the help of Whitman filter paper. Or study included total 18 patients out of which there 12 females and 6 males with the overall ocular frequency 33.3%. In Uveitis cases, there were 7 females and 9 males with overall percentage 29.6%. Scleritis had 6 females and 6 males with cumulative frequency of 22.2%. While peripheral keratitis was present in 4 females and 2 males with overall frequency of 11.1%. The age group categorization of the study included most patients of autoimmune rheumatic cases in between 36 to 45 years. Late adult onset has some striking association with hormonal changes. Keratoconjunctivitis siccas were predominant in between the age of 27-36 years whereas uveitis and scleritis were found in predominance 36-45 years. Our study also postulated the linkage between the steroids and ocular manifestations, as 5 of the patients had steroid-onset ocular diseases and all those cases were of Keratoconjunctivitis Sicca.

### Conclusion

To summarize, in our retrospective cross-sectional study, the most prevalent rheumatic autoimmune disease was Rheumatoid Arthritis (RA). And the resultant ocular complication was Keratoconjunctivitis Sicca. In view of the studies done on the linkage between the rheumatic autoimmune diseases and the ocular diseases in Pakistan as well as in other countries, our study showed an overall decrease in the ocular involvement.

## RHEUMATIC AUTOIMMUNE DISEASE

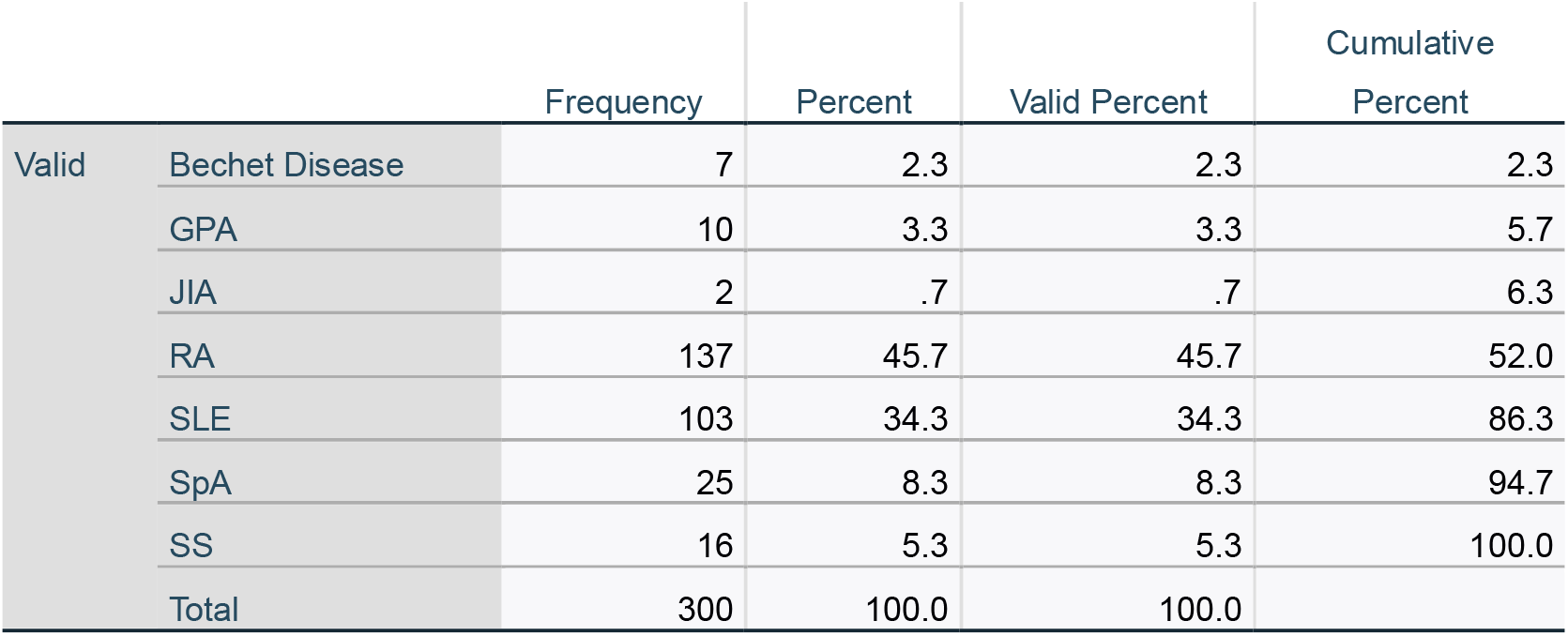

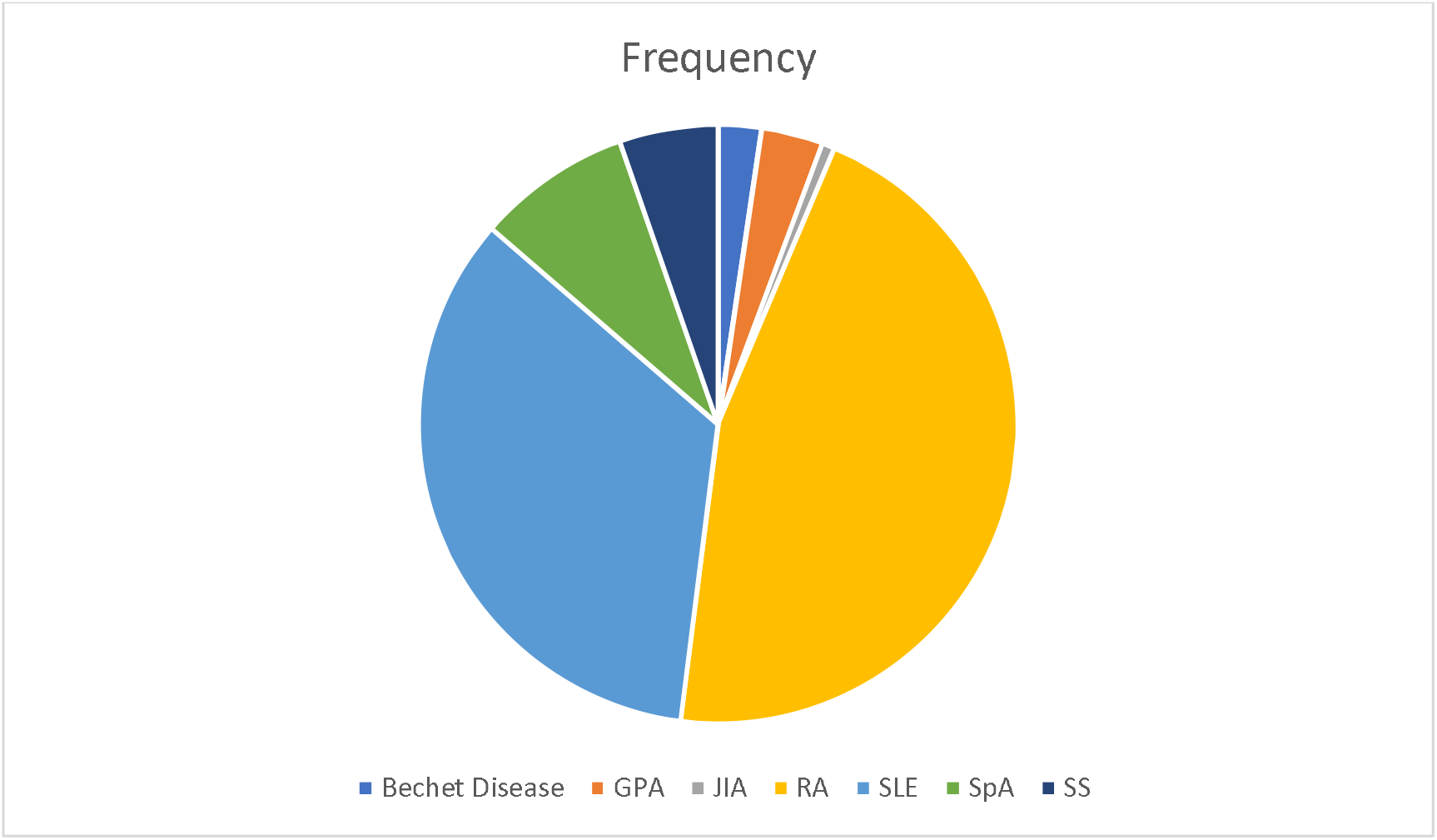

### SEX * RAD Crosstabulation

**SEX \* RAD Crosstabulation:**
|  | Bechet Disease | GPA | JIA | RA | SLE | SpA | SS | Total |
| --- | --- | --- | --- | --- | --- | --- | --- | --- |
| Female | 7 | 4 | 0 | 87 | 69 | 2 | 10 | 179 |
| Male | 0 | 6 | 2 | 50 | 34 | 23 | 6 | 121 |
| Total | 7 | 10 | 2 | 137 | 103 | 25 | 16 | 300 |

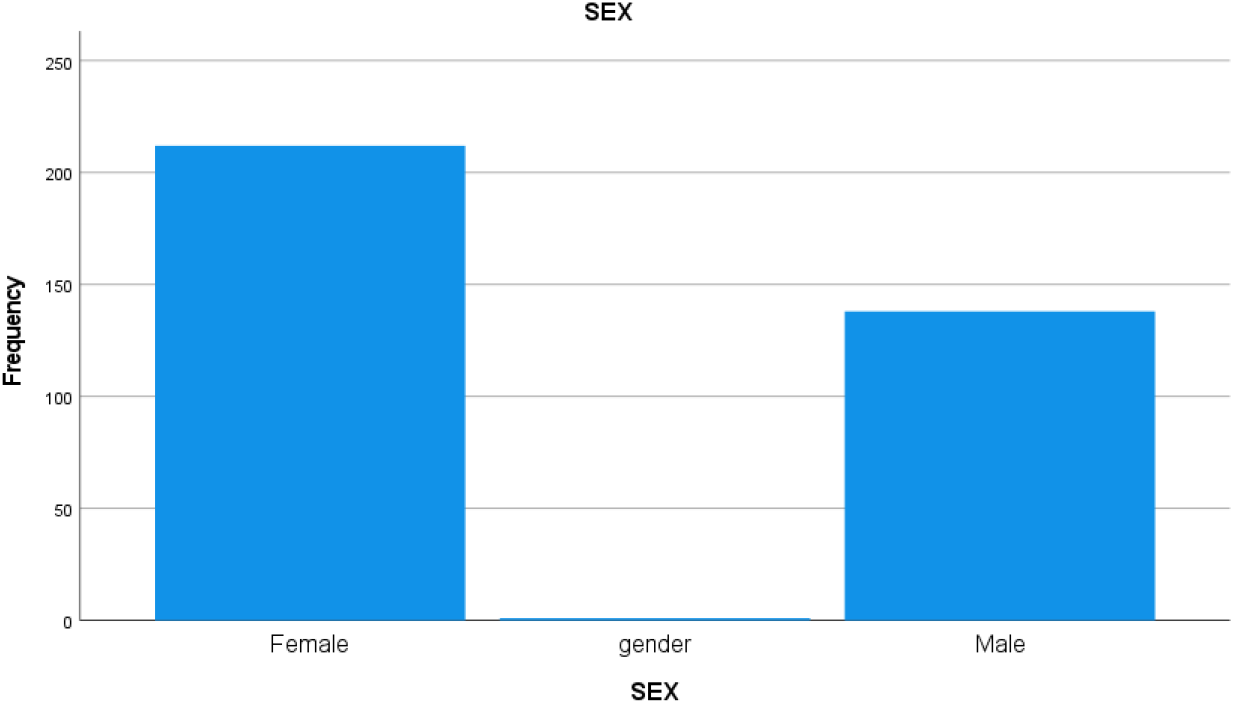

## DISTRIBUTION ACCORDING TO AGE

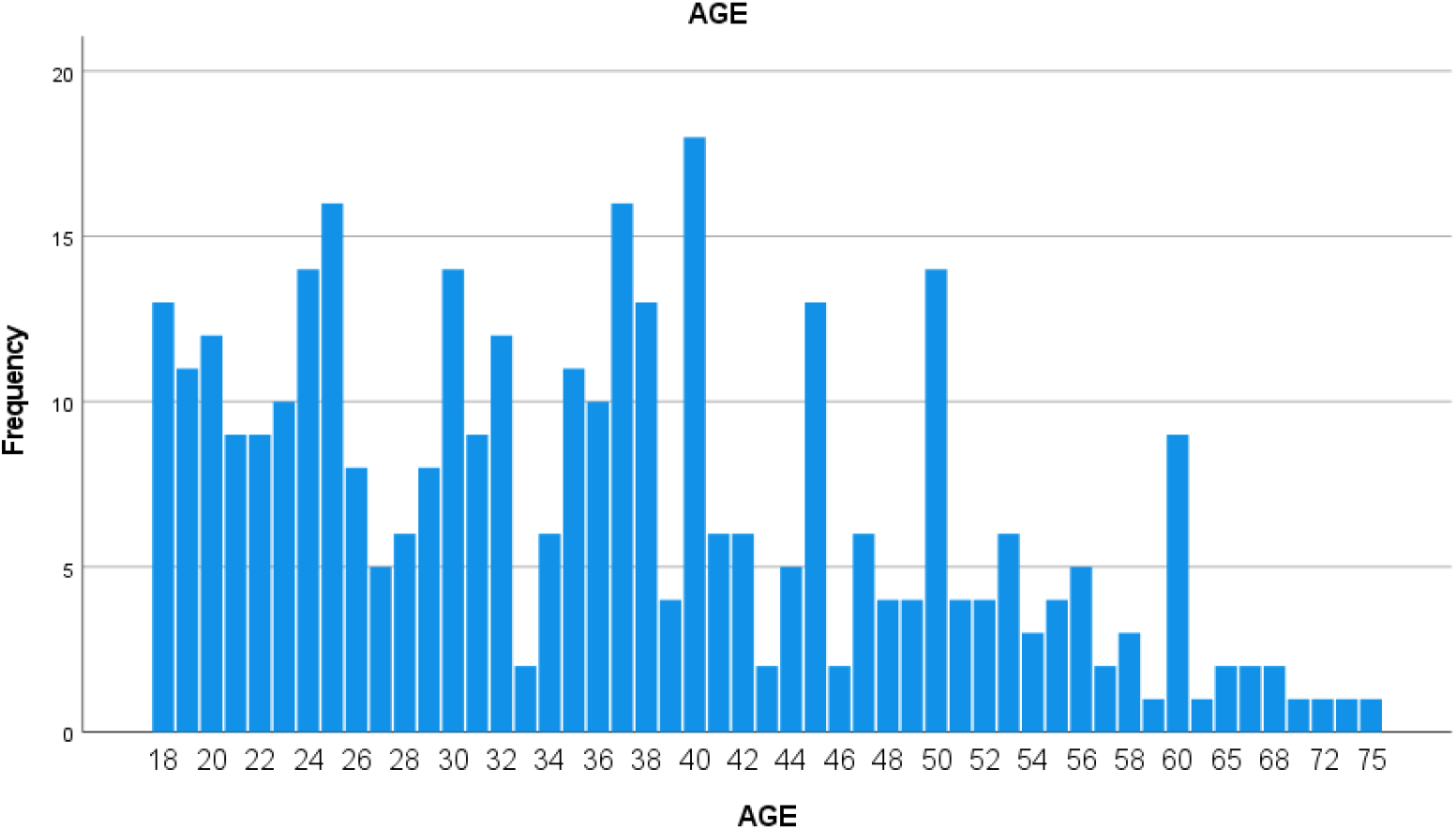

## OCCULAR DISEASES

**OCCULAR DISEASES:**
|  | Frequency | Percent | Valid Percent | Commulative Percent |
| --- | --- | --- | --- | --- |
| Keratoconjunctivitis Sicca | 18 | 6.0 | 6.0 | 88.0 |
| Peripheral Ulcer Keratitis | 6 | 2.0 | 2.0 | 90.0 |
| Retinal Vasculitis | 2 | .7 | .7 | 90.7 |
| Scleritis | 12 | 4.0 | 4.0 | 94.7 |
| Uveitis | 16 | 5.3 | 5.3 | 100.0 |
| Total | 300 | 100.0 | 100.0 |  |

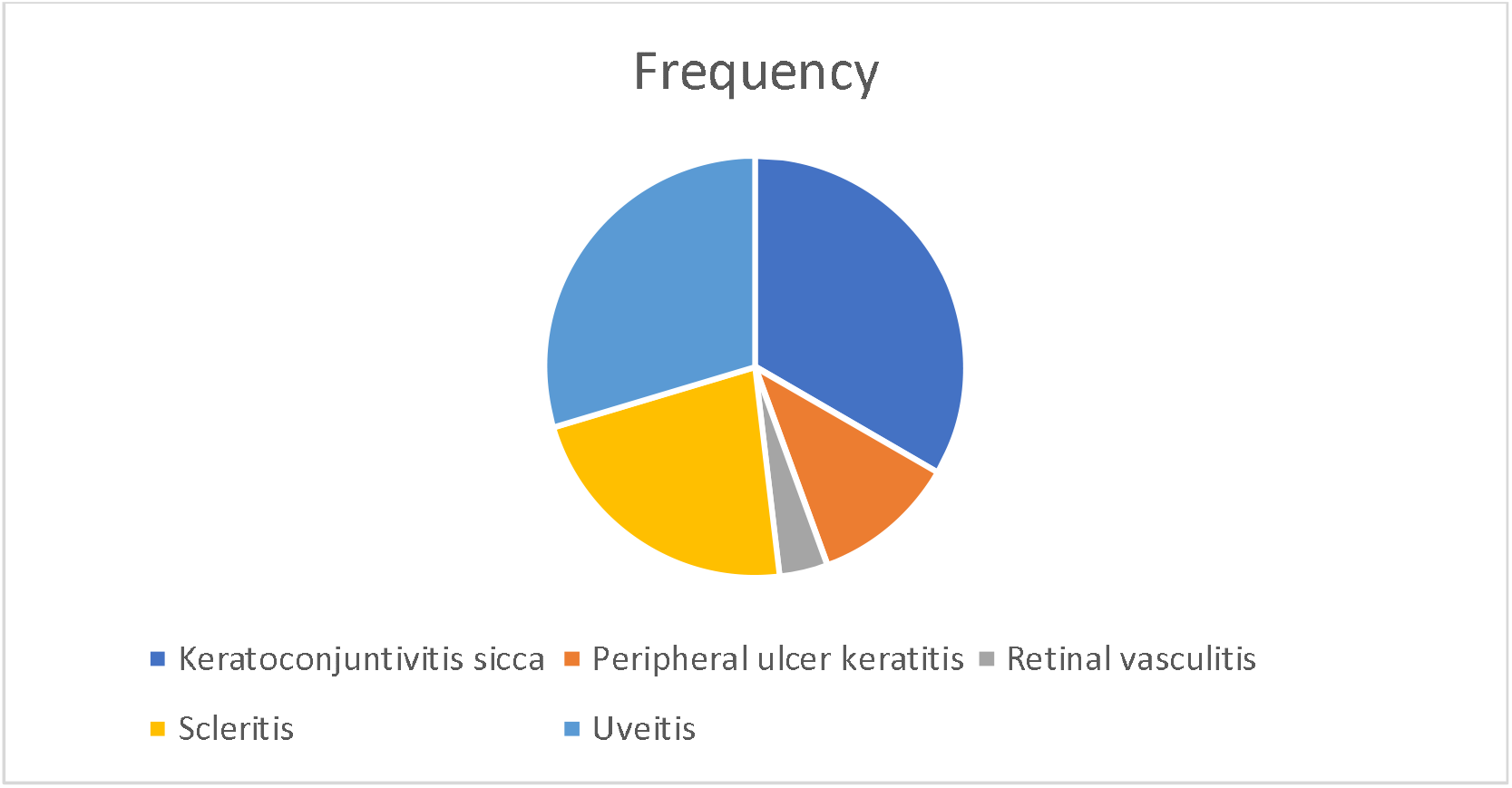

## Notes

### Competing Interest Statement

The authors have declared no competing interest.

### Funding Statement

This study did not receive any funding.

### Author Declarations

Ethics committee of Shaikh Zayed Hospital Lahore gave ethical approval for this work. As my work includes collection of data from people suffering from ocular manifestations with autoimmune rheumatic disorders so the ethics committee gave ethical approval for this work.

